# Omicron Subvariants: Clinical, Laboratory, and Cell Culture Characterization

**DOI:** 10.1101/2022.09.20.22280154

**Authors:** C. Paul Morris, Raghda E. Eldesouki, Jaiprasath Sachithanandham, Amary Fall, Julie M. Norton, Omar Abdullah, Nicholas Gallagher, Maggie Li, Andrew Pekosz, Eili Y. Klein, Heba H. Mostafa

## Abstract

**Background:** The variant of concern, Omicron, has become the sole circulating SARS-CoV-2 variant for the past several months. Omicron subvariants BA.1, BA.2, BA.3, BA.4, and BA.5 evolved over the time, with BA.1 causing the largest wave of infections globally in December 2021- January 2022. In this study, we compare the clinical outcomes in patients infected with different Omicron subvariants and compare the relative viral loads, and recovery of infectious virus from upper respiratory specimens.

**Methods:** SARS-CoV-2 positive remnant clinical specimens, diagnosed at the Johns Hopkins Microbiology Laboratory between December 2021 and July 2022, were used for whole genome sequencing. The clinical outcomes of infections with Omicron subvariants were compared to infections with BA.1. Cycle threshold values (Ct) and the recovery of infectious virus on VeroTMPRSS2 cell line from clinical specimens were compared.

**Results:** The BA.1 was associated with the largest increase in SARS-CoV-2 positivity rate and COVID-19 related hospitalizations at the Johns Hopkins system. After a peak in January cases fell in the spring, but the emergence of BA.2.12.1 followed by BA.5 in May 2022 led to an increase in case positivity and admissions. BA.1 infections had a lower mean Ct when compared to other Omicron subvariants. BA.5 samples had a greater likelihood of having infectious virus at Ct values less than 20.

**Conclusions:** Omicron subvariants continue to associate with a relatively high positivity and admissions. The BA.5 infections are more while BA.2 infections are less likely to have infectious virus, suggesting potential differences in infectibility during the Omicron waves.

**Funding:** Centers for Disease Control and Prevention contract 75D30121C11061, NIH/NIAID Center of Excellence in Influenza Research and Surveillance contract HHS N2772201400007C, Johns Hopkins University, Maryland department of health, and The Modeling Infectious Diseases in Healthcare Network (MInD) under awards U01CK000589.

## Introduction

Multiple subvariants of Omicron have emerged since its first discovery in November 2021 (1, 2). In the United States, BA.1 predominated in December 2021 and January 2022 then was displaced by BA.2, followed by BA.2.12.1 in March and April of 2022. The BA.4 and BA.5 then displaced all other Omicron sublineages, with BA.5 becoming dominant despite having an identical Spike protein sequence to BA.4 (https://covid.cdc.gov/covid-data-tracker/#variant-proportions last accessed 8/22/22). The evolution of each subvariant was associated with increasing evasion of neutralizing antibodies. The BA.1 and BA.2 showed a large reduction in neutralization by antibodies induced by vaccination, prior infection, as well as therapeutic monoclonal antibodies (3-6). The BA.2.12.1 and the BA.4/BA.5 showed increased neutralization escape compared to BA.2 (7, 8).

The Omicron variants were first discovered in South Africa and Botswana in November 2021, however, the kinetics of reporting of its subvariants were ahead in this region compared to the United States (9). The South African experience revealed lower number of cases, hospital admissions, and deaths during the BA.4/BA.5 wave when compared to the BA.1 wave (9, 10), even though BA.4/BA.5 caused a large number of reinfections (10). As the behavior of Omicron subvariants might be impacted by the differences in levels of immunity to prior infections and vaccination rates which are both significantly different in the US compared to South Africa, in this study we examined the outcomes of infection with Omicron subvariants for patients diagnosed at the Johns Hopkins system. In addition, we provide a comparison of upper respiratory viral loads from patients infected with the Omicron subvariants and the recovery of infectious virus on cell culture.

## Methods

### Ethical considerations and Data availability

The research was performed with a waiver of consent under the Johns Hopkins protocol IRB00221396. Whole genomes that met the quality standards were made publicly available at GISAID.

### Specimens and Patients’ Data

In this retrospective observational cohort study, we used nasopharyngeal or lateral mid-turbinate nasal swabs from remnant clinical specimens from the Johns Hopkins Health System (JHHS) after standard of care SARS-CoV-2 diagnostic or screening testing for symptomatic and asymptomatic patients. Specimens included samples obtained from across all inpatient and outpatient settings in the National Capital Region (4, 11). SARS-CoV-2 testing was performed using different molecular approaches as we described before (4, 6, 11-14).

### Sample size

We included all Omicron infections identified from November 25^th^, 2021, the date the first Omicron variant identified in our system (11), through July 17^th^, 2022. Because only 3 Omicron infections were identified in our system in November, total infections and positivity rates were calculated from the beginning of December 2021. For patients who were tested more than once, we used their initial collection. Whole genome sequencing for surveillance was attempted for all positive specimens with left-over volumes from JHHS and samples are collected in real-time on a daily basis. All samples with genomes that did not pass a quality of coverage > 90% and mean depth >100 were excluded. Genomes with unassigned lineages were excluded. For Ct and cell culture experiments, samples were randomly selected from the whole cohort based on availability (Table 1).

**Table 1.** Patients and samples used for the study.

| **lineage** | **BA.1** | **BA.1.1** | **BA.2** | **BA.2.12.1** | **BA.4** | **BA.5** |
| --- | --- | --- | --- | --- | --- | --- |
| Total patients | 3334 | 637 | 1041 | 1267 | 191 | 484 |
| Total with complete clinical data | 3285 | 637 | 1038 | 1234 | 166 | 337 |
| Unvaccinated (%) | 1114 (33.9) | 219 (34.4) | 372 (35.8) | 510 (41.3) | 73 (44) | 124 (36.8) |
| Fully Vaccinated (%) | 1448 (44.1) | 240 (37.7) | 242 (23.3) | 256 (20.7) | 29 (17.5) | 85 (25.2) |
| Boosted (%) | 723 (22) | 178 (27.9) | 424 (40.8) | 468 (37.9) | 64 (38.6) | 128 (38) |
| **Cell culture** |  |  |  |  |  |  |
| Cell culture (Total tested) | 239 | 27 | 166 | 68 | 88 | 125 |
| Cell culture (% positive) | 63.60 | 48.15 | 27.71 | 41.18 | 56.82 | 60.00 |
| Unvaccinated (% positive) | 69.74 | 60.00 | 23.26 | 36.36 | 53.85 | 61.11 |
| Vaccinated (% positive) | 64.84 | 55.56 | 30.95 | 35.00 | 73.33 | 56.41 |
| Boosted (% positive) | 55.56 | 25.00 | 28.40 | 50.00 | 52.94 | 62.00 |
| **Ct values** |  |  |  |  |  |  |
| Ct values (Total tested) | 1695 | 515 | 785 | 868 | 154 | 417 |
| Ct values (Mean) | 19.4 | 22.8 | 22.7 | 22.21 | 22.5 | 21.9 |
| Ct values (SD) | 4.9 | 5.5 | 5.9 | 5.4 | 5.4 | 5.3 |
| Less than 5 days (total) | 1013 | 284 | 416 | 484 | 75 | 173 |
| Asymptomatic (total) | 229 | 101 | 127 | 107 | 17 | 35 |
| Unvaccinated | 555 | 176 | 277 | 352 | 63 | 110 |
| Vaccinated | 776 | 195 | 174 | 182 | 24 | 80 |
| Boosted | 364 | 144 | 332 | 309 | 50 | 112 |
| **Disposition** |  |  |  |  |  |  |
| ICU (%) | 1.4 | 2.2 | 1.6 | 1.9 | 1.8 | 1.8 |
| ICU (Total) | 45 | 14 | 17 | 24 | 3 | 6 |
| Inpatient (%) | 4.4 | 5.2 | 7.5 | 8.7 | 9.6 | 10.4 |
| Inpatient (Total) | 145 | 33.0 | 78.0 | 107.0 | 16.0 | 35.0 |
| Immunosuppressed (%) | 17.1 | 14.0 | 15.6 | 17.8 | 17.4 | 17.2 |
| Immunosuppressed (Total) | 561.0 | 89.0 | 162.0 | 220.0 | 29.0 | 58.0 |
| Mean patients age | 37.7 | 37.9 | 39.7 | 38.7 | 36.2 | 43.6 |

### Clinical data analysis

Clinical and vaccination data for patients whose samples were characterized by whole genome sequencing was bulk extracted as previously detailed (4, 11) and cases with missing data were excluded. Because every patient admitted to a JHHS hospital is tested for SARS-CoV-2 regardless of symptoms, we used presenting complaints, ED admission diagnoses, reason for testing, and timing of testing relative to admission to distinguish between patients who were admitted primarily for COVID-19 treatment from incidental asymptomatic admissions. Patients with an ED diagnosis related to an upper respiratory infection or with chief complaints consistent with COVID-19 (based on influenza like illness (ILI) syndromic surveillance) or whose reason for testing was other than asymptomatic, were considered a COVID-related admission. Patients tested under asymptomatic admission protocols were considered non-COVID-related admissions. Each admission was scored based on the likelihood of the admission being COVID-related based on the complaints, diagnoses, and reasons for testing. Full vaccination was based on positive SARS-CoV-2 test results more than 14 days after the second shot for pfizer/BioNTech BNT162b2 and Moderna mRNA-1273 or 14 days after the J&J/Janssen. In our vaccinated patients’ population, 65.1% received Pfizer/BioNTech, Moderna mRNA-1273 (30.1%), and the J&J/Janssen COVID-19 vaccines (4.8%).

### Ct value analysis

To ensure comparable Ct values for relative viral load analyses, Ct values of the N gene were collected. For that, samples were retested with either the PerkinElmers SARS-CoV-2 kit (https://www.fda.gov/media/136410/download, Last accessed August 17, 2022) or the CDC designed primers and probes for the N gene (14).

### Amplicon based Sequencing

Specimen preparation, extractions, and sequencing were performed as described previously (11, 15, 16). Library preparation was performed using the NEBNext® ARTIC SARS-CoV-2 Companion Kit (VarSkip Short SARS-CoV-2 # E7660-L). Sequencing was performed using the Nanopore GridION and analysis was performed as described in (11). Sequences with coverage >90% and mean depth >100 were submitted to GISAID database. Genomes with lineages assigned by Pangolin were included (coverage > 70%, Tables S1 details the quality of the genomes).

### Cell culture

VeroE6TMPRSS2 (VT) cells (RRID: CVCL_YQ49) were obtained from the National Institute of Infectious Diseases, Japan and are described in (5) and were processed as we described previously (11). VeroE6-ACE2-TMPRSS2 (VAT) cells were obtained from the BEI resources repository and cultured in an identical manner to VT cells. The cells were cultured and infected with aliquots of swab specimens as previously described for VeroE6 cells (17) except that 75 μL was added to VT or VAT cells for experiments that assessed parallel virus isolations. Cultures were incubated for 7 days or until cytopathic effect (CPE) was obvious and SARS-CoV-2 infection was confirmed by reverse transcriptase PCR (17). Wells with approximately 75% of the cells showing CPE were considered positive and the day this occurred was documented.

### Viruses

SARS-CoV-2/USA-WA1/2020 (EPI_ISL_404895) was obtained from BEI Resources. The other SARS-CoV-2 viruses used in this study were hCoV19/USA/MD-HP00076/2020 (EPI_ISL_438234), hCoV19/USA/MD-HP11011/2021 (EPI_ISL_825013), hCoV19/USA/CA-VRLC088/2021 (EPI_ISL_2987142), hCoV19/USA/MD-HP07626/2021 (EPI_ISL_3373222), hCoV19/USA/MD-HP05660/2021 (EPI_ISL_2331507), hCoV19/USA/MD-HP25001/2022 (EPI_ISL_9245416), hCoV19/USA/MD-HP28972/2022 (EPI_ISL_11962964), hCoV19/USA/MD-HP32103/2022 (GISAID accession number pending) and hCoV19/USA/MD-HP30386/2022 (EPI_ISL_12416220), which were all isolated from COVID-19 patients at JHHS as previously described (11).

### Tissue culture infective dose (TCID_50_) assay for infectious SARS-CoV-2 titer quantification

The infectious virus titer was determined on VT or VAT cells using a 50% tissue culture infectious dose (TCID_50_) assay as previously described for SARS-CoV-2 (11). VT or VAT cells were grown to 90-100% confluence in 96 well plates. Serial 10-fold dilutions of the virus stock were made in infection media (IM) (identical to CM except the FBS was reduced to 2.5%), and then 20 μL of each dilution was added to the cells in sextuplicate. The cells were incubated at 37°C with 5% CO_2_ for 5 days. The cells were fixed by adding 100 μL of 4% formaldehyde in PBS per well overnight and visualized by staining with 75 μL of naphthol blue-black solution overnight and scored visually for cytopathic effect. A Reed and Muench calculation were used to determine the TCID50 per mL.

### Statistical analysis

Chi-square analysis was used for categorical variables with and without correction for confounding variables. For association between lineage and hospitalization or mortality, age and vaccine status were controlled using Cochran-Mantel-Haenszel method utilizing 5 categories for continuous variables (3). Where appropriate, Fisher Exact test was used for categorical variable comparisons. One-way ANOVA and t test were used for comparing continuous independent variables. Post-hoc analysis was carried out with Mann-Whitney U test with Bonferroni correction where appropriate.

## Results

### SARS-CoV-2 positivity and Omicron subvariants trends December 2021-July 2022

The monthly SARS-CoV-2 positivity rate was highest in January 2022 (23.7%) then declined to a mean of 1.6% in March 2022 before increasing again in May 2022 to a mean of 7.6% (Figure 1A). The positivity rate then largely plateaued through July 2022 (Figure 1A). The predominant Omicron subvariant in December 2021 and January 2022 was BA.1 (82.7% and 96.3% respectively, Figure 1B). Other subvariants displaced the BA.1 including the BA.1.1 in February (58%), BA.2 in March and April (52.2% and 66.9%), the BA.2.12.1 in May and June (53.6% and 49.5%), and the BA.5 in July (62.9%). Notably, COVID-19 related admissions peaked in January 2022, correlating with the peak of BA.1 then declined in March and April, before increasing again in May 2022 and plateauing similar to the positivity rate (Figure 1C and D).

**Figure 1.**
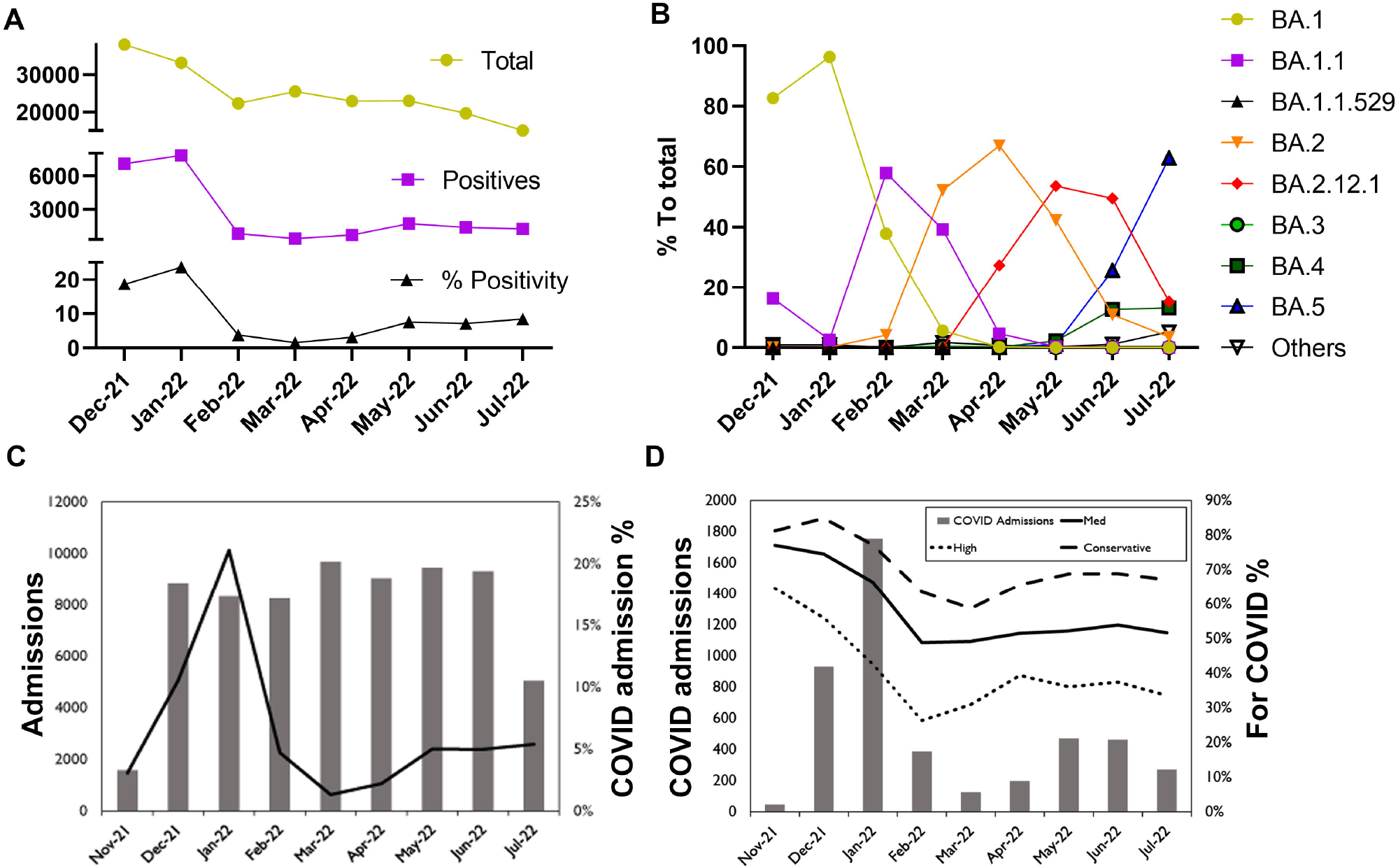
SARS-CoV-2 positivity, Omicron subvariants trends, and COVID-19 related admissions, December 2021-July 17, 2022. A) SARS-CoV-2 average monthly total tests, total positives, and positivity rates for both symptomatic and asymptomatic testing. B) SARS-CoV-2 Omicron subvariant average monthly percent to total sequenced. C) Total patients admitted each month as inpatients (includes hospitalized observations) in JHHS between November 25^th^, 2021 and July 17, 2022 (excludes neonates born in the hospital). COVID-19 hospitalizations includes any patient with a positive test in the 14 days prior to admission or within the first 48 hours. D) Total patients admitted with a positive COVID-19 test in the 14 days prior to admission or within the 48 hours after admission. “For COVID” is an estimate of the percentage of patients that were admitted because of COVID-19 and not for a different issue then had an incidental positive laboratory test with no symptoms during hospital screening. The high case only includes patients flagged as having symptoms and not flagged as asymptomatic and had influenza like illness (ILI) complaints and an ILI diagnosis at admission. The med case includes all high cases as well as cases that only had an ILI complaint or ILI diagnosis or noted symptoms at admission and were not tested asymptomatically. The conservative case included cases were there was either an ILI complaint or ILI diagnosis or noted symptoms at admission but were tested with an asymptomatic flag or there was no ILI or symptoms noted but there was not an asymptomatic flag on the test.

### Patient characteristics and outcomes in infections caused by Omicron subvariants

Of 199,378 samples tested for SARS-CoV-2 from JHHS between November 25th 2021 and July 17th 2022, a total of 21,007 samples were positive, of them, 11,775 were sequenced for genomic surveillance and 8,377 were of high quality. After excluding repeat tests in patients and all other clades than Omicron, virus genome sequencing identified a total of 6,993 unique patients who were infected with Omicron subvariants. Only the major subvariants that showed high prevalence were used for further analysis (N = 6,954, Table 1). Subvariants BA.1.1, BA.2, BA.2.12.1, BA.4, and BA.5 were compared to subvariant BA.1 (which we characterized as compared to Delta in a prior study (11)). Samples within branches from subvariants were traced back to lineages used in the study (i.e. BA.1.2 would be considered BA.1, and BA.1.1.2 would be considered BA.1.1, Table S1). Notably, the percentages of infections in individuals who received booster vaccination increased from 22% with BA.1 to 40.8%, 37.9%, 38.6%, and 38% with BA.2, BA.2.12.1, BA.4, and BA.5 (Table 1).

Compared to BA.1, the odds ratio for COVID-19 related hospitalization was higher with BA.1.1 (1.2, p = 0.45), BA.2 (1.57, p = 0.0032), BA.2.12.1 (1.62, p = 0.001), BA.4 (1.87, p = 0.023), and BA.5 (1.66, p = 0.021) after controlling for patients’ age and vaccine status with lower likelihood of COVID-19 related mortality (Table 2). Similar trends were observed when these variables were not accounted for (Table 3). In general, fully vaccinated patients and those who received booster vaccination were less likely to be hospitalized (0.83 (p = 0.09) and 0.92 (p = 0.47)), and different underlying conditions increased the likelihood of admissions including primarily kidney disease, heart disease, and immune suppression (Table 3).

**Table 2.** Odds ratios of Omicron subvariants related hospitalization compared to BA.1.

| Age/vaccine-controlled odds ratio<br>(reference BA.1) | Hospitalization | P value compared<br>to BA.1 | Mortality | P value compared<br>to BA.1 |
| --- | --- | --- | --- | --- |
| BA.1.1 | 1.2 | 0.45 | 0.76 | 0.64 |
| BA.2 | 1.57 | 0.003 | 0.57 | 0.26 |
| BA.2.12.1 | 1.62 | 0.001 | 0.81 | 0.57 |
| BA.4 | 1.87 | 0.023 | 0.95 | 0.95 |
| BA.5 | 1.66 | 0.021 | 0.22 | 0.081 |

**Table 3.** Odds of Omicron related admissions in our study cohort.

| **Variable** | **Odds** | **p** |
| --- | --- | --- |
| **Lineage** |  |  |
| BA.1 | 0.54 | <0.001 |
| BA.1.1 | 0.81 | 0.300 |
| BA.2 | 1.28 | 0.059 |
| BA.2.12.1 | 1.59 | <0.001 |
| BA.4 | 1.64 | 0.071 |
| BA.5 | 1.83 | 0.002 |
| **Vaccine status** |  |  |
| Vaccinated | 0.83 | 0.09 |
| Boosted | 0.92 | 0.47 |
| **Comorbidities** |  |  |
| Hypertension | 5.22 | <0.001 |
| Pregnancy | 4.03 | 0.01 |
| Lung Disease | 1.97 | <0.001 |
| Kidney Disease | 9.00 | <0.001 |
| Immunosuppression | 7.10 | <0.001 |
| Diabetes | 4.90 | <0.001 |
| Heart Failure | 8.29 | <0.001 |
| Atrial Fibrillation | 5.83 | <0.001 |
| Smoker | 2.52 | <0.001 |
| Cerebrovascular Disease | 5.07 | <0.001 |
| Cancer | 2.05 | <0.001 |
| Coronary Artery Disease | 7.41 | <0.001 |

### Omicron subvariants cycle threshold (Ct) values in upper respiratory samples

The mean Ct value of BA.1 samples (19.43) was significantly lower compared to all other subvariants (BA.1.1 (22.81), BA.2 (22.74), BA.2.12.1 (22.74), BA.4 (22.56), BA.5 (21.92), one-way ANOVA, p < 0.0001, Figure 2A). This held true when comparing Ct values from symptomatic cases within the first 5 days of symptoms (BA.1 (18.79), BA.1.1 (22.26), BA.2 (22.06), BA.2.12.1 (21.76), BA.4 (22.39), BA.5 (21.41), one-way ANOVA, p < 0.0001, Figure 2B), and when comparing Ct values from asymptomatic patients (BA.1 (21.23), BA.1.1 (24.42), BA.2 (24.24), BA.2.12.1 (23.79), BA.4 (23.7), BA.5 (24.74), one-way ANOVA, p < 0.0001, Figure 2C). No differences were noted when Ct values were compared between vaccinated and unvaccinated patients from all groups, however, the mean Ct of BA.1 was consistently lower than the other subvariants in unvaccinated, fully vaccinated, and patients who received a booster vaccination (Figure 2D). Figure 2E and Table 1 and 4 summarize the numbers tested for each subvariant.

**Table 4.** Cycle thresholds (Ct) total, mean, and standard deviations (SD) for Omicron subvariants by vaccination status.

|  |  | Ct value (N gene) |  |  |
| --- | --- | --- | --- | --- |
|  |  | count | mean | SD |
| lineage | Vaccine status |  |  |  |
| BA.1 | Unvaccinated | 555 | 19.84244 | 4.841083 |
|  | Vaccinated | 776 | 19.24888 | 4.929083 |
|  | Boosted | 365 | 19.19809 | 4.847137 |
| BA.1.1 | Unvaccinated | 176 | 22.63063 | 5.857535 |
|  | Vaccinated | 195 | 23.10762 | 5.296218 |
|  | Boosted | 144 | 22.63257 | 5.192578 |
| BA.2 | Unvaccinated | 277 | 22.66242 | 6.125257 |
|  | Vaccinated | 174 | 22.87009 | 6.122025 |
|  | Boosted | 332 | 22.7156 | 5.70073 |
| BA.2.12.1 | Unvaccinated | 352 | 22.41742 | 5.555199 |
|  | Vaccinated | 182 | 22.25526 | 5.336134 |
|  | Boosted | 309 | 22.11507 | 5.525919 |
| BA.4 | Unvaccinated | 63 | 22.05164 | 5.868287 |
|  | Vaccinated | 24 | 22.41408 | 5.155246 |
|  | Boosted | 50 | 23.39513 | 5.253949 |
| BA.5 | Unvaccinated | 110 | 22.00935 | 5.182746 |
|  | Vaccinated | 80 | 23.06307 | 6.267613 |
|  | Boosted | 112 | 21.76022 | 4.993582 |

**Figure 2.**
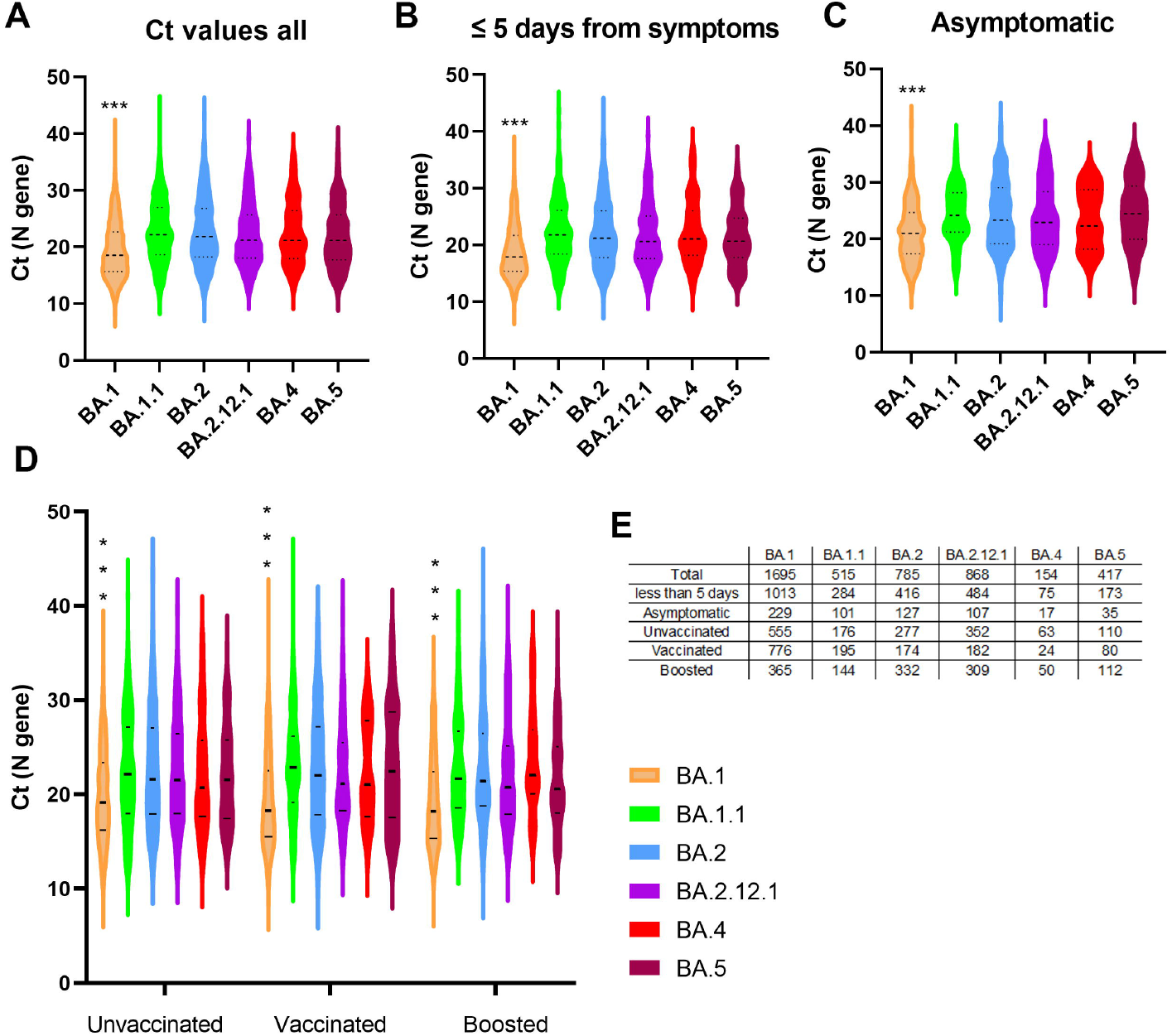
Omicron subvariants cycle threshold (Ct) values in upper respiratory samples. A) Ct values of Omicron subvariants from all samples with available Ct values (N gene). B) Ct values of Omicron subvariants from samples collected 5 days or less from the onset of symptoms. For this analysis, samples from asymptomatic patients were not included. C) Ct values of Omicron subvariants collected from patients with “asymptomatc” status in the charts. D) Ct values from Omicron subvariants groups stratified by vaccination status. E) Total samples used for each Omicron subvariant. Vaccinated, fully vaccinated patients who didn’t receive a booster dose; boosted, patients with booster dose. Data shown as violin plots and horizontal bars represent medians and quartiles.

### Recovery of infectious virus in Omicron subvariant groups

Recovery of infectious virus was performed for 713 samples on VT cells (Table 1 and 5), the same cell line that we used to compare Omicron to Delta in a prior study (11). The recovery of infectious virus was higher from samples with lower mean Ct values regardless of the subvariant (Figure 3A, mean Ct for positive versus negative cell culture: BA.1 (16.4 vs 20.5, p < 0.0001), BA.1.1 (18.5 vs 25.1, p = 0.004), BA.2 (20.3 vs 24.9, p < 0.0001), BA.2.12.1 (21.2 vs 23.7, p = 0.19), BA.4 (19.3 vs 25.1, p = 0.0001), BA.5 (18.5 vs 26.4, p < 0.0001). Recovery rates for Omicron subvariants were not significantly different from BA.1, with the exception of BA.2 and BA.2.12.1 which consistently showed less recovery of infectious virus on VT cells, regardless of patients’ vaccination status, when the relative virus loads of the samples were not accounted for (Figure 3B and Table 5). When we used samples with Ct values less than 20 to compare between groups, BA.5 showed statistically higher recovery of infectious virus (Figure 3C and Table 5).

**Table 5.** Cell culture results of Omicron subvariants (VT cell line). Two tailed P values were calculated by Fisher Exact test.

| **Cell culture** | **BA.1** | **BA.1.1** | **BA.2** | **BA.2.12.1** | **BA.4** | **BA.5** |
| --- | --- | --- | --- | --- | --- | --- |
| Total | 239 | 27 | 166 | 68 | 88 | 125 |
| negative | 87 | 14 | 120 | 40 | 38 | 50 |
| positive | 152 | 13 | 46 | 28 | 50 | 75 |
| P value |  | 0.14 | 0.0001 | 0.0013 | 0.3 | 0.57 |
| **Unvaccinated** | 76 | 10 | 43 | 22 | 39 | 36 |
| negative | 23 | 4 | 33 | 14 | 18 | 14 |
| positive | 53 | 6 | 10 | 8 | 21 | 22 |
| P value |  | 0.7 | 0.0001 | 0.006 | 0.1 | 0.39 |
| **Vaccinated** | 91 | 9 | 42 | 20 | 15 | 39 |
| negative | 32 | 4 | 29 | 13 | 4 | 17 |
| positive | 59 | 5 | 13 | 7 | 11 | 22 |
| P value |  | 0.7 | 0.0003 | 0.022 | 0.77 | 0.43 |
| **Boosted** | 72 | 8 | 81 | 26 | 34 | 50 |
| negatives | 32 | 6 | 58 | 13 | 16 | 19 |
| positive | 40 | 2 | 23 | 13 | 18 | 31 |
| P value |  | 0.14 | 0.0009 | 0.65 | 0.83 | 0.58 |
| **Ct < 20 by vaccination status** |  |  |  |  |  |  |
| **Unvaccinated** | 45 | 3 | 13 | 8 | 13 | 15 |
| negatives | 9 | 0 | 8 | 3 | 3 | 1 |
| positives | 36 | 3 | 5 | 5 | 10 | 14 |
| P value |  | 1 | 0.01 | 0.36 | 1 | 0.43 |
| **Vaccinated** | 63 | 3 | 16 | 4 | 4 | 13 |
| negative | 16 | 0 | 9 | 2 | 0 | 0 |
| positive | 47 | 3 | 7 | 2 | 4 | 13 |
| P value |  | 1 | 0.032 | 0.29 | 0.56 | 0.058 |
| **Boosted** | 38 | 3 | 22 | 3 | 6 | 17 |
| negatives | 13 | 2 | 10 | 0 | 1 | 1 |
| positive | 25 | 1 | 12 | 3 | 5 | 16 |
| P value |  | 0.54 | 0.42 | 0.54 | 0.65 | 0.042 |
| **Total less than 20** | 146 | 9 | 51 | 15 | 23 | 45 |
| negatives | 38 | 2 | 27 | 5 | 4 | 2 |
| positives | 108 | 7 | 24 | 10 | 19 | 43 |
| P value |  | 1 | 0.0009 | 0.55 | 0.45 | 0.001 |

**Figure 3.**
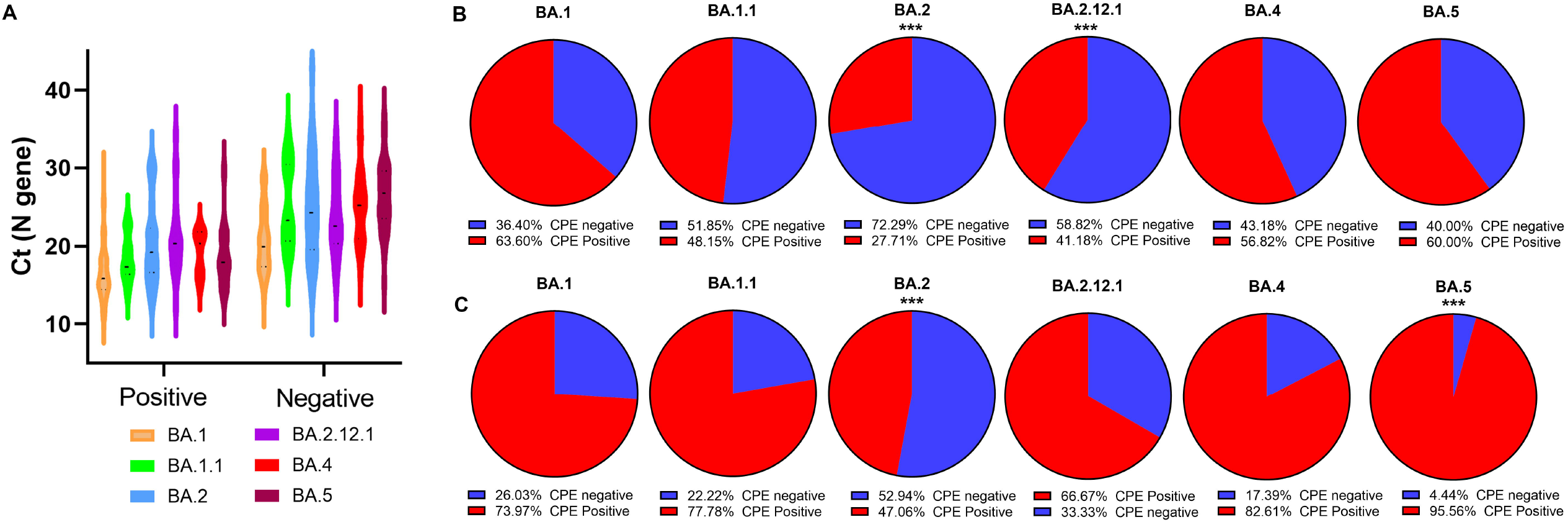
Recovery of infectious virus from respiratory samples of patients infected with Omicron subvariants. A) Total positives and negatives for Omicron subvariants in association with Ct values. B) Percent positives for the recovery of infectious virus with Omicron subvariants for all Ct values or for samples with Ct values less than 20 (C). CPE: cytopathic effect.

### Sensitivity of VAT versus VT cell lines for the recovery of infectious Omicron subvariants

To assess the difference of the recovery of infectious Omicron subvariants if we use a cell line that expresses ACE-2 in addition to TMPRSS2, 332 samples were cultured side by side on both VAT and VT cell lines. A significant increase in the recovery of infectious virus was notable on VAT compared to VT for all samples (59.3% vs 47.3%, P = 0.0031, Figure 4A), as well as for samples with Ct more than 20 (45.3% vs 31.3%, P = 0.012, Figure 4B), but the difference did not reach statistical significance for samples with Ct < 20 (Figure 4C and Table 6). Only BA.5 samples with Ct more than 20 showed a significant difference in virus recovery between VAT and VT (Table 6).

**Table 6.** Cell culture results of Omicron subvariants on VT versus VAT cell lines. Two tailed P values were calculated by Fisher Exact test.

| **Cell culture** | **BA.1** | **BA.1.1** | **BA.2** | **BA.2.12.1** | **BA.4** | **BA.5** | **All subvariants** |
| --- | --- | --- | --- | --- | --- | --- | --- |
| **Total** | 8 | 27 | 65 | 66 | 88 | 78 | 332 |
| **VT** |  |  |  |  |  |  |  |
| negative | 7 | 14 | 45 | 39 | 38 | 31 | 174 |
| positive | 1 | 13 | 20 | 27 | 50 | 47 | 158 |
| **VAT** |  |  |  |  |  |  |  |
| negative | 5 | 11 | 35 | 32 | 32 | 20 | 135 |
| positive | 3 | 16 | 30 | 34 | 56 | 58 | 197 |
| P value | 0.57 | 0.59 | 0.1 | 0.29 | 0.44 | 0.09 | 0.0031 |
| **Ct < 20** |  |  |  |  |  |  |  |
| **Total** | 2 | 9 | 25 | 15 | 23 | 28 | 102 |
| **VT** |  |  |  |  |  |  |  |
| negative | 1 | 2 | 13 | 5 | 4 | 2 | 27 |
| positive | 1 | 7 | 12 | 10 | 19 | 26 | 75 |
| **VAT** |  |  |  |  |  |  |  |
| negative | 1 | 2 | 9 | 2 | 2 | 2 | 18 |
| positive | 1 | 7 | 16 | 13 | 21 | 26 | 84 |
| P value | 1 | 1 | 0.39 | 0.39 | 0.67 | 1 | 0.18 |
| **Ct > 20** |  |  |  |  |  |  |  |
| **Total** | 5 | 10 | 31 | 34 | 47 | 36 | 163 |
| **VT** |  |  |  |  |  |  |  |
| negative | 5 | 8 | 24 | 21 | 29 | 25 | 112 |
| positive | 0 | 2 | 7 | 13 | 18 | 11 | 51 |
| **VAT** |  |  |  |  |  |  |  |
| negative | 3 | 6 | 20 | 20 | 25 | 15 | 89 |
| positive | 2 | 4 | 11 | 14 | 22 | 21 | 74 |
| P value | 0.44 | 0.63 | 0.4 | 1 | 0.5 | 0.032 | 0.012 |

**Figure 4.**
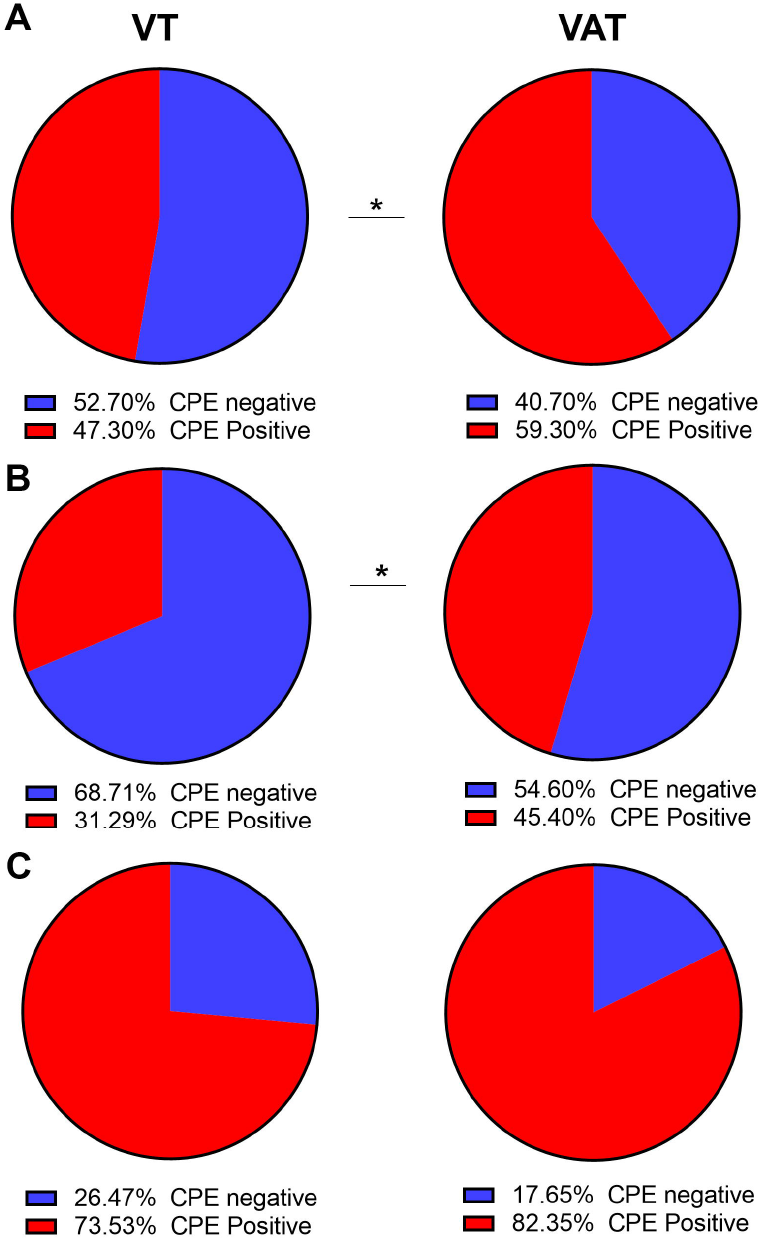
Recovery of infectious virus from respiratory samples of patients infected with Omicron subvariants on VT versus VAT cell lines. A) Percent positives and negatives for Omicron subvariants. B) Percent positives and negatives for samples with Ct values more than 20 (C) Percent positives and negatives for Omicron subvariants with Ct values less than 20. CPE: cytopathic effect.

To further assess if VAT cell line enhances the isolation of Omicron subvariants, we selected 4 different Omicron subvariants’ isolates and compared their TCID_50_ on VAT versus VT cell lines. We included the wild type strain (WAS, SARS-CoV-2/USA-WA1/2020) in addition to some pre-Omicron lineages (Table 7). The average log TCID_50_ of 4 replicates per each isolate was consistently higher and mostly statistically significant when VAT cells were used with an average log difference of 0.47 between VAT and VT cell lines (Table 7).

**Table 7.**
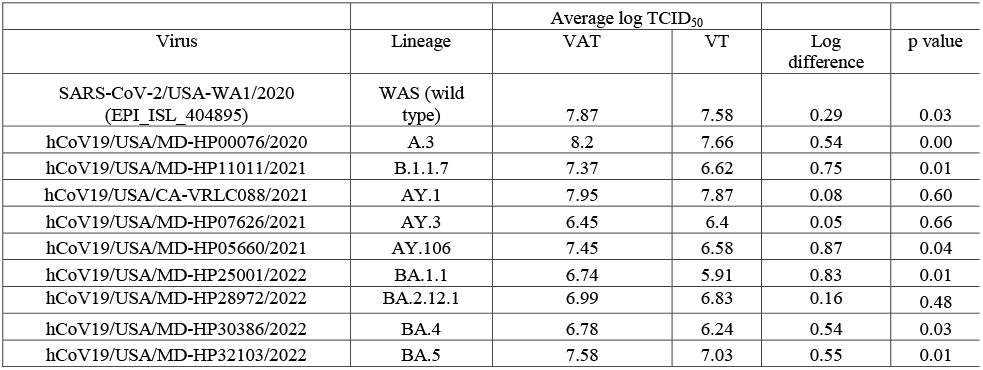
TCID_50_ results of select Omicron subvariants on VT versus VAT cell lines. Two tailed P values were calculated by *t* test.

### Reinfections caused by Omicron subvariants

To assess the possibility of reinfections with Omicron subvariants after a prior infection with Omicron, we evaluated the timing of multiple positives from the same patients in our cohort. There were 170 likely reinfections with lineages BA.2, BA.2.12.1, BA.4 or BA.5 based on patients having multiple positive samples with an initial positive occurring prior to the first detection of that lineage. Median days from initial sample to reinfection was 167 days (mean of 236). For samples with an initial infection in December 2021 or January 2022, which was likely BA.1 or BA.1.1, a total of 133 were identified as reinfected with another Omicron subvariant (Table 8).

**Table 8.** Reinfections caused by Omicron subvariants with an initial infection after December 1^st^ (Omicron primary infection).

|  |  | First known positive sample |  |
| --- | --- | --- | --- |
|  |  | Initial positive after<br>December 1st 2021 | Total<br>reinfections |
| lineage | Vaccine status |  |  |
| **BA.2** | Boosted | 24 | 27 |
|  | Unvaccinated | 15 | 18 |
|  | Vaccinated | 9 | 13 |
| **BA.2.12.1** | Boosted | 20 | 31 |
|  | Unvaccinated | 21 | 26 |
|  | Vaccinated | 9 | 14 |
| **BA.4** | Boosted | 7 | 7 |
|  | Unvaccinated | 6 | 7 |
|  | Vaccinated | 3 | 3 |
| **BA.5** | Boosted | 9 | 10 |
|  | Unvaccinated | 7 | 10 |
|  | Vaccinated | 3 | 4 |

## Discussion

In this study, a large retrospective cohort of patients infected with Omicron between December 2021 (in addition to three infected patients in November 2021) and July 2022 was used to compare outcomes of infection by the most predominant subvariants. Our data showed that the largest peak of SARS-CoV-2 positivity and COVID-19 related admissions was in December 2021 and January 2022 and associated with the BA.1 wave. Subsequent predominant Omicron subvariants included the BA.1.1 in February 2022, BA.2 in March and April 2022, BA.2.12.1 in May and June 2022, and BA.5 in July 2022. Those waves correlated with a reduction in cases and admissions in February to April 2022, followed by a small but plateaued increase in May to July 2022. Comparing COVID-19 related admissions for each lineage, showed that there was a slight increase in the likelihood of admission in BA.1.1, BA.2, BA.2.12.1, BA.4, and BA.5 compared to BA.1 but a reduction in the likelihood of mortality. Admissions were more likely in patients with different comorbidities.

Despite the increased likelihood in admission of all subvariants studied compared to BA.1, this does not necessarily mean that the other variants can inherently cause a more severe disease than BA.1. As a study in South Africa did not report a difference in hospitalization rates with BA.2 compared to BA.1 (18), it seems likely that other factors could contribute to increased admissions in our cohort. The differences in hospitalization rates for each lineage in our study could reflect stricter criteria for admission during elevated rates of infection, increased testing at home leading to only more serious cases being captured by our screening methods, or seasonality, in which colder and drier months are associated with more COVID-19 cases (19) as well as a potentially waning immune response leading to more severe respiratory infections.

Infections with BA.1 were associated with the large increase in the number of cases and hospitalization in December 2021 and in January 2022 (11). Even though, BA.2 displaced BA.1, its predominance did not correlate with an increase in the number of cases; on the contrary, in our system, when BA.2 was predominant, it was associated with a period of the least positivity rates since the emergence of Omicron (March and April, 2022). Interestingly, our cell culture model showed a reduction in the recovery of infectious virus from BA.2 samples which might reflect a reduction in infectivity of BA.2 in our region. Remarkably, this was not a consistent pattern worldwide, and countries that included Denmark, China, and Japan reported an increase in hospitalizations and death during the peak of BA.2 (20-22). This indicates that the emergence, spread, and risk of different subvariants are likely dependent on many factors within a community that include the immune responses due to prior infections or vaccinations. Several reports showed that booster vaccination or prior COVID-19 vaccination followed by SARS-CoV-2 infection are associated with an increase in the neutralizing antibodies to the Omicron subvariants (7, 23). This might explain the differences between countries that had a massive circulation of BA.1 followed by the emergence of BA.2 and other Omicron subvariants versus other countries that experienced probably the co-circulation of BA.1 and BA.2.

In our study, we show that BA.1 infections were associated with the highest relative virus load in upper respiratory specimens when compared to the subsequent Omicron subvariants. In a previous study, when we compared the relative upper respiratory viral loads in BA.1 and Delta infected patients, we didn’t notice a significant difference between the two variants (11). Groups from Tokyo and France did not report a significant difference in viral loads between BA.1 and BA.2 infected cases (24, 25). The data indicate that the selective advantage of these subvariants is likely not due to higher upper respiratory viral loads. The discrepancy between our findings and prior reports might emphasize the impact of the community level immune landscape that very likely differ by geographical locations.

It is puzzling that, in spite of the identical spike regions of the BA.4 and BA.5, the BA.5 had greater success in community transmission and has become predominant, even though both variants were first detected around the same time in our geographical region. Both variants also showed a marked reduction in neutralization by antibodies. We also show that in our cohort, both BA.4 and BA.5 were capable of causing re-infections in patients who likely had a prior infection with BA.1 or BA.1.1. In our study, we show that the BA.5 is associated with more recovery of infectious virus on cell culture when we controlled for the specimens’ viral load. Outside of spike protein, the BA.4 and BA.5 differ in other regions that include the ORF7b:L11F, N:P151S, and deletions NSP1:141–143 in BA.4 and M:D3N change in BA.5 (9), in addition to a significant divergence in the 3’ end. It was shown before that Omicron has higher affinity to ACE-2 than Delta and that its spike uses TMPRSS2 inefficiently (26). When we compared the recovery of infectious virus from different Omicron subvariants on VT versus VAT, we noticed an increased sensitivity with VAT that was more notable for samples with lower relative viral loads. Interestingly, an overexpression of ACE-2 was advantageous for not only Omicron subvariants, but also to the original SARS-CoV-2 and pre-Omicron variants as shown by our TCID_50_ comparison and consistent with prior reports (27). Further characterization of these two Omicron subvariants on cell culture might reveal the genomic basis of the increased BA.5 viral fitness noted in our study.

We previously showed that the recovery of infectious virus on cell culture inversely correlates with SARS-CoV-2 specific IgG in respiratory specimens (4, 11, 28). We also showed that Omicron was associated with the largest peak of reinfections in our system (29). The reduction in the recovery of infectious virus from BA.2 specimens and the increased recovery of infectious virus from BA.5 specimens might be due to differences in the neutralization of these subvariants in upper respiratory specimens by SARS-CoV-2 specific antibodies against a vaccine spike or a prior infection. We believe that immune escape contributes to the increased infectivity and peaks of increased positivity and reinfections.

This study has many limitations. First, information and samples were limited to patients tested within JHHS. This means that this study lacks a true knowledge of the number of cases occurring at any time, or the true number of cases in the community for a particular lineage. Additionally, information on admissions elsewhere and patients vaccinated in other locations/states may not always be documented within our system. Lastly, testing did not occur at the same interval from the start of symptoms which can impact the viral load or ability to recover culturable virus from samples. While we used time from symptoms to normalize the groups being tested for Ct values, time from the start of symptoms may not fully represent the time from the start of infection and is subject to recall bias and documentation challenges.

## Data Availability

All data produced in the present study are available upon reasonable request to the authors.

## Acknowledgements

This study was only possible with the unique efforts of the Johns Hopkins Clinical Microbiology Laboratory faculty and staff. HHM is supported by the HIV Prevention Trials Network (HPTN) sponsored by the National Institute of Allergy and Infectious Diseases (NIAID). Funding was provided by the Centers for Disease Control (contract 75D30121C11061), the Johns Hopkins Center of Excellence in Influenza Research and Surveillance (HHSN272201400007C), National Institute on Drug Abuse, National Institute of Mental Health, and Office of AIDS Research, of the NIH, DHHS (UM1 AI068613), the NIH RADx-Tech program (3U54HL143541-02S2), National Institute of Health RADx-UP initiative (Grant R01 DA045556-04S1), the Johns Hopkins University President’s Fund Research Response, the Johns Hopkins department of Pathology, and the Maryland department of health. EK was supported by Centers for Disease Control and Prevention (CDC) MInD-Healthcare Program (Grant Number U01CK000589). The views expressed in this manuscript are those of the authors and do not necessarily represent the views of the National Institute of Biomedical Imaging and Bioengineering; the National Heart, Lung, and Blood Institute; the National Institutes of Health, or the U.S. Department of Health and Human Services.

## Data sharing

Whole genome data were made available publicly (GISAID IDs, Table S1) and raw genomic data requests could be directed to HHM.

